# Early genomic, epidemiological, and clinical description of the SARS-CoV-2 Omicron variant in Mexico City

**DOI:** 10.1101/2022.02.06.22270482

**Authors:** Alberto Cedro-Tanda, Laura Gómez-Romero, Guillermo de Anda-Jauregui, Dora Garnica-López, Yair Alfaro-Mora, Sonia Sánchez-Xochipa, F. Eulices García-García, Alfredo Mendoza-Vargas, J. Emmanuel Frías-Jiménez, Bernardo Moreno-Quiroga, Abraham Campos-Romero, José Luis Moreno-Camacho, Jonathan Alcantar-Fernández, Jesús Ortíz-Ramírez, Mariana Benitez-Gonzalez, Roxana Trejo-Gonzalez, Daniel Aguirre-Chavarria, Marcela E. Núñez-Martínez, Laura Uribe-Figueroa, Ofelia Angulo, Rosaura Ruiz, Alfredo Hidalgo-Miranda, Luis A Herrera

**Affiliations:** Instituto Nacional de Medicina Genómica, Mexico City, Mexico; Researchers for Mexico (previously Cátedras CONACYT para jóvenes investigadores), Mexico City, Mexico; Centro de Ciencias de la Complejidad, UNAM, Mexico City, Mexico; Innovation and Research Department, Salud Digna, Sinaloa, Mexico; Clinical Laboratory Division, Salud Digna, Sinaloa, Mexico; Hospital General Ajusco Medio, SEDESA, Mexico City, Mexico; Centro Médico ABC, Mexico City, Mexico; Laboratorio Arion Genética, Mexico City, Mexico; Secretaría de Educación, Ciencia, Tecnología e Innovación de la Ciudad de México; Unidad de Investigación Biomédica en Cáncer, Instituto Nacional de Cancerología-Instituto de Investigaciones Biomédicas, UNAM

## Abstract

**Background:** Omicron is the most mutated SARS-CoV-2 variant that has emerged, resulting in viral phenotype alterations, which can affect transmissibility, disease severity, and immune evasiveness. Genomic surveillance of a highly transmissible variant is important in cities with millions of inhabitants and an economic center such as Mexico City. In this work, we describe the early effects of the Omicron variant in Mexico City, exploring its genomic profile and clinical description.

**Methodology:** We sequenced SARS-CoV-2-positive samples in November and December 2021 and we using the public database GISAID. Haplotype and phylogenetic analyses were performed to genomically characterize Omicron. We used the Mexican federal database to explore the association with clinical information such as symptoms and vaccination status.

**Findings:** The first case of Omicron was detected on November 16, 2022, and until December 31, 2021, we observed an increase from 88% in sequenced samples. Nineteen nonsynonymous mutations were found in the Omicron RBD, and we further explored the R346K substitution, which was prevalent in 42% of the samples and associated with immune escape by monoclonal antibodies. In the phylogenetic analysis, we found that there were several independent exchanges between Mexico and the world, and there was an event followed by local transmission that gave rise to most of the Omicron diversity in Mexico City. The haplotype analysis allowed us to observe that there was no association between haplotype and vaccination status. Of the patients with clinical data, 66% were vaccinated, none of the reported comorbidities were associated with Omicron, the presence of odynophagia and absence of dysgeusia were significant predictor symptoms for Omicron, and the Ct value on RT–qPCR was lower in Omicron.

**Conclusions:** Genomic surveillance in highly populated and fast-moving urban regions such as Mexico City is key to detecting the emergence and spread of SARS-CoV-2 variants in a timely manner, even weeks before the onset of an infection wave, to detect patterns that can inform public health decisions. It is also necessary to continue sequencing to detect the spread of any mutation that may affect the therapeutic efficacy or guide it.

## Introduction

Despite the great efforts made by the worldwide scientific and health community to contain the COVID-19 pandemic, a major challenge has been the emergence of novel variants of concern (VOCs) of SARS-CoV-2. These variants exhibit new features, such as increased transmissibility, immune escape, and other adaptations, that make epidemic control more difficult [1] The latest of these variants of concern is B.1.1.529, also known as the Omicron variant.

The first world case of the Omicron variant of SARS-CoV-2 was identified in South Africa on 2 November 2021, rapidly spreading to 104 countries on 13 January 2022 [2]. The Omicron variant has rapidly become the dominant variant worldwide. Omicron has the highest number of mutations compared with the other VOCs; these mutations have been associated with increased transmissibility, resistance to therapy, and partially escaping immunity induced by infections or vaccines [3].

Mexico City has been heavily affected by the COVID-19 pandemic. Two intense epidemic waves occurred in spring 2020 and winter 2020/2021, during which the heaviest toll of hospitalizations and deaths was observed [4,5]. A vaccination campaign began, starting with frontline healthcare workers and continuing in descending age order, and was completed in fall 2021 (http://vacunacovid.gob.mx/wordpress/calendario-vacunacion/). In summer 2021, a third wave, associated with the delta variant, occurred, with reduced case hospitalization and fatality rates. After a period of downward epidemic trends, the first case associated with the Omicron variant was detected on November 16, 2021, in Mexico City, preceding an ongoing fourth wave.

As the capital and largest city in the country, the behavior of the pandemic in Mexico City has a large impact on the national epidemic patterns. As the economic and transportation hub of the country (https://datamexico.org/), cases can be easily imported and exported to other regions throughout the country. Furthermore, having one of the most important international airports in the country, it can be a major entry point for new variants from abroad [6]. For these reasons, the control of SARS-CoV-2 spread in Mexico City is paramount to protect both its local population and its connationals throughout the country. In this regard, genomic surveillance is a major tool to identify and assess novel threats [7]. Early detection of the importation and spread of new variants of concern, complementing the monitoring of epidemic and public health metrics, may allow policymakers to adapt and rapidly implement strategies to mitigate the epidemic phenomenon.

In this work, we present an analysis of the early effects of the Omicron variant in Mexico City during its first month of spread. We identify the likely importation pattern, with several importation events followed by local transmission. We show that the growth of the epidemic curve is associated with an increased prevalence of the Omicron variant, at the expense of the previously dominant delta variant. We also show the high prevalence of the R346K mutation. We show the emergence of different haplotypes during this period. We study differences in clinical presentation with the coexisting delta variant, finding little evidence of neither lessened nor increased severity. We discuss the implications of these insights for policymaking, highlighting the importance of genomic surveillance associated with epidemiological monitoring in the context of the current pandemic.

## Materials and methods

### Participants

Nasopharyngeal swabs (NPS) were collected from patients for SARS-CoV-2 detection. The study was approved by the ethic and research committee of the Instituto Nacional de Medicina Genómica (CEI/1479/20 and CEI 2020/21), all procedures were in accordance with the ethical standards of the institutional research committee.

### Sample collection

NPSs were collected by a trained clinician with a flexible nylon swab that was inserted into the patient’s nostrils to reach the posterior nasopharynx. It was left in place for several seconds and slowly removed while rotating. The swab was then placed in 2 mL of sterile viral transport medium. Swabs from both nostrils were deposited in a single viral transport tube, taken to a clinical laboratory and processed immediately.

### SARS-CoV-2 RNA extraction

Total nucleic acid was extracted from 300 µL of viral transport medium from the NPSs or 300 µL of whole saliva using the MagMAX Viral/Pathogen Nucleic Acid Isolation Kit (Thermo Fisher Scientific, Waltham, MA, USA) and eluted into 50 µL of elution buffer.

### RT–qPCR detection

For SARS-CoV-2 RNA detection, 5 µL of RNA template was tested using TaqPath master mix (Thermo Fisher Scientific, Waltham, MA, USA). All tests were run on a Thermo Fisher ABI QuantStudio 5 real-time thermal cycler (Thermo Fisher Scientific, Waltham, MA, USA). Samples were selected for inclusion in this study based on viral Ct < 30.

### Sequencing sample

Illumina Sequencing. The libraries were prepared using the Illumina COVID-seq protocol following the manufacturer’s instructions. First-strand synthesis was carried out on RNA samples. The synthesized cDNA was amplified using ARTIC primers V3 for multiplex PCR, generating 98 amplicons across the SARS-CoV-2 genome. The PCR-amplified product was tagmented and adapted using IDT for Illumina Nextera UD Indices Set A, B, C, D (384 indices) (Illumina, San Diego, CA, USA). Dual-indexed pair-end sequencing with a 36 bp read length was carried out on the NextSeq 550 platform (Illumina, San Diego, CA, USA). 2.2.4. Illumina Raw Data Processing and Sequencing Data Quality Assessment The raw data were processed using DRAGEN Lineage v3.3.4/.5 with standard parameters (Illumina, San Diego, CA, USA). Further samples with SARS-CoV-2 and at least 90 targets detected were processed for lineage designation.

### Clinical data acquisition

We used the COVID-19 database hosted by the federal health authorities to explore different demographic and clinical information. Briefly, this database contains clinical information captured during an initial interview, including age, sex, 20 symptoms, 10 comorbidities, date of symptom onset, hospitalization status, death date (if applicable), and vaccination date. Since the Mexican vaccination strategy considers the use of several different vaccines, for the purposes of this work, we consider a patient to be fully vaccinated 14 days after the last dose specified for the vaccine used (monodose or two-dose regimes).

Sequenced data were matched to clinical information using an internal lab id. All cases from samples with symptom onset from epidemiological weeks 2021-48 to 2021-52 identified in medical units in Mexico City were kept. Samples not belonging to the delta and Omicron variants of interest (VOC) were discarded.

### Comparisons between delta and Omicron populations

We performed statistical analyses to assess whether the delta and Omicron populations exhibited differences in any variable of interest. We used a combination of statistical tests based on the comparisons of interest:

- For numerical variables (age, number of comorbidities, number of symptoms), we used a two-sided t test.
- For categorical variables, we used Fisher’s exact test to test for overrepresentation of the feature (equivalent to difference in proportions) within any of the variants.
- For multivariate analyses, we performed logistic regressions of the form

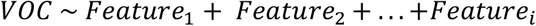

All statistical analyses were performed using R version 4.1.2 (2021-11-01) -- “Bird Hippie.” Data manipulation and visualization were performed using Tidyverse tools [8], and statistical tests were performed using the Tidymodels suite.

### Genetic background description

SARS-CoV-2 Omicron genomes from Mexico City were downloaded from GISAID [9], with the following criteria: data of sample collection from November 14 to December 31, 2021, only complete sequences with a genome coverage >95% (n=783), submission until January 25, 2021. The Nextclade tool was used to determine amino acid substitutions in the viral genome and generate histogram plots of mutation frequency and phylogenies highlighting mutation occurrence [10].

### Variant calling and haplotype analysis

Sequences of Omicron SARS-CoV-2 genomes from Mexico City were downloaded from GISAID. Only complete sequences with an N fraction lower than 0.05 were considered (N = 136). The SARS-CoV-2 reference genome NC_045512.2 was downloaded from NCBI. SNVs per SARS-CoV-2 sequence were obtained with nucmer [11]Nucmer was executed with the following parameters: map each position of each query to its best hit in the reference, map each position of each reference to its best hit in the query and exclude alignments with ambiguous mapping. Variable positions in any SARS-CoV-2 sequence were obtained. Only variable positions observed in at least 13 genomes were further considered. Each SARS-CoV-2 sequence was translated into a compressed representation including only the genotype of the variable positions. A unique combination of alleles, e.g., a unique compressed representation, was considered a haplotype. Haplotypes were used to infer a haplotype network using the haploNet function from the Population and Evolutionary Genetics Analysis System package (pegas) [12]. Briefly, genetic distances (Hamming distance) between all pairwise combinations of haplotypes were calculated using the dist.dna function of the Analyses of Phylogenetics and Evolution package (ape) [13] from this distance matrix, and the minimum spanning tree and the median-joining network were computed using pegas [12]. The number of sequences per haplotype was obtained. The most ancient sequence per haplotype was defined as the one with the most ancient date of onset of symptoms.

### Phylogenetic analysis

We downloaded the full Multiple Sequence Alignment from Gisaid on January 11, 2022. This MSA was performed based on 6,900,922 submissions to EpiCoV. The full methods are described on the Gisaid website. Briefly, both duplicate and low-quality sequences (>5% Ns) were removed using only complete sequences (length >29,000 bp). Each sequence was individually aligned to the reference hCoV-19/Wuhan/WIV04/2019, discarding dubious alignments. Sequences that result in unique insertions in the reference sequence and that occurred more than once are used as an initial set of sequences for multiple sequence alignment, reducing each contiguous stretch of NNNs into a single letter N. The remaining sequences are aligned to the resulting alignment (este es el resumen de abajo). From this MSA, we calculated the Hamming distance from any sequence of our collection to any of the MSA sequences. We selected the closest sequences (Hamming distance <= 10). We calculated the best-scoring Maximum Likelihood tree using the model GTR + Optimization of substitution rates + Optimization of site-specific evolutionary rates (50 bootstrap analysis) via raxmlHPC [14] We used iTOL for tree visualization [15].

## Results and discussion

### Phylogenetic analysis

We performed a phylogenetic analysis to infer the possible origin of Omicron SARS-CoV-2 in Mexico. We analyzed all Omicron SARS-CoV-2 collected in Mexico together with their closest worldwide relatives. The best-scoring maximum likelihood (ML) phylogeny is presented in Figure 1. We can observe that several exchange events between Mexico and the rest of the world have occurred since sequences collected in Mexico are located all over the phylogeny. We can also observe that several independent exchange events have occurred between Mexico and the USA (black arrows). However, the direction of these exchange events cannot be interpreted from this analysis. Additionally, several exchange events cannot be associated with a specific country. For example, Figure 1B shows a monophyletic group formed by sequences from Mexico, United States of America (USA), United Kingdom (UK), France, Germany and Ecuador, and Figure 1C (lower clade) shows a monophyletic group formed by sequences from Germany, Israel, USA, Mexico, France and UK. Interestingly, most exchange events between Mexico and the rest of the world are associated with groups of sequences that present a low distance to their foreign counterparts; however, there is one group of sequences collected in Mexico that have largely diverged from the rest of the world (gray arrow), displaying shorter distances between them compared to any other distances. The origin of this monophyletic group of sequences is unclear. Interestingly, most of the exchange events have associated sequences from Mexico City and from the rest of the country (Figure 1D); however, the most divergent monophyletic group contains sequences from only Mexico City (gray arrow). The observed pattern suggests multiple independent exchange events between Mexico and the rest of the world in the first month of Omicron occurrence in Mexico. Concurrently, it also suggests a single event followed by local transmission that gave rise to most of the diversity of Omicron SARS-CoV-2 sequences observed in Mexico City. The scenario depicted in Mexico is not rare since independent importations of specific SARS-CoV-2 variants have been described in the literature [16,17] as well as superspreading events following recurrent importations [18] This behavior can be associated with frequent human movement between and within country borders.

**Figure 1.**
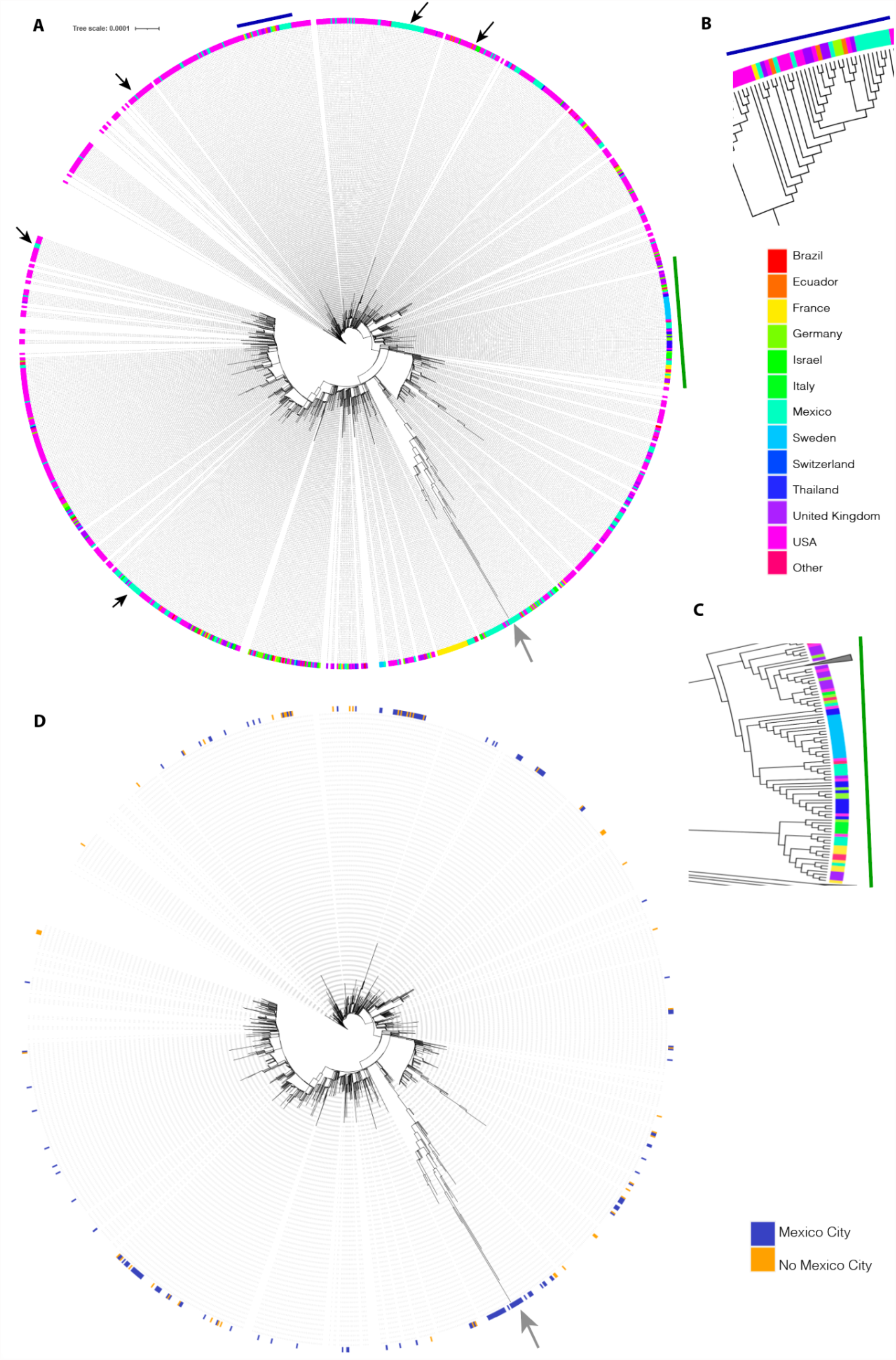
The best-scoring ML phylogeny of Omicron SARS-CoV-2 collected in Mexico together with their closest worldwide relatives. Panel A. The best-scoring ML phylogeny of all sequences is presented. Black arrows indicate exchange events between the USA and Mexico. The gray arrow displays a group of sequences collected in Mexico that has largely diverged from the rest of the sequences. Panel B. A zoom on the region indicated by the blue line. Panel C. A zoom on the region indicated by the green line. Colors represent countries in which the analyzed sequences were collected. Panel D. Colors represent blue, sequences collected in Mexico City; orange, sequences collected in Mexico but not in Mexico City.

### Identification of Omicron variant in Mexico City

The first case of the Omicron variant in Mexico was detected on November 16, 2021, in Mexico City, and as of December 31, 2021, 783 cases of Omicron were detected by next-generation sequencing in Mexico City. According to the pangolin lineage, 57% were BA.1 and 43% BA.1.1 [19].

At week 46, the Omicron variant represented 1.85%, while the Delta variant represented 98.15%. Then, in week 50, the Omicron variant increased rapidly, reaching 65% and 35% for Delta. At week 52, the Omicron variant was 88%, and the delta variant was 12% (Figure 2A and 2B). At the same time, (week 52), the Omicron variant had different prevalences in the Americas, e.g., USA 59.29%, Brazil 37.57%, Colombia 60.37% and Argentina 45.8%, while in the UK it was 69.33%, and 98.87% in South Africa [2].

**Figure 2.**
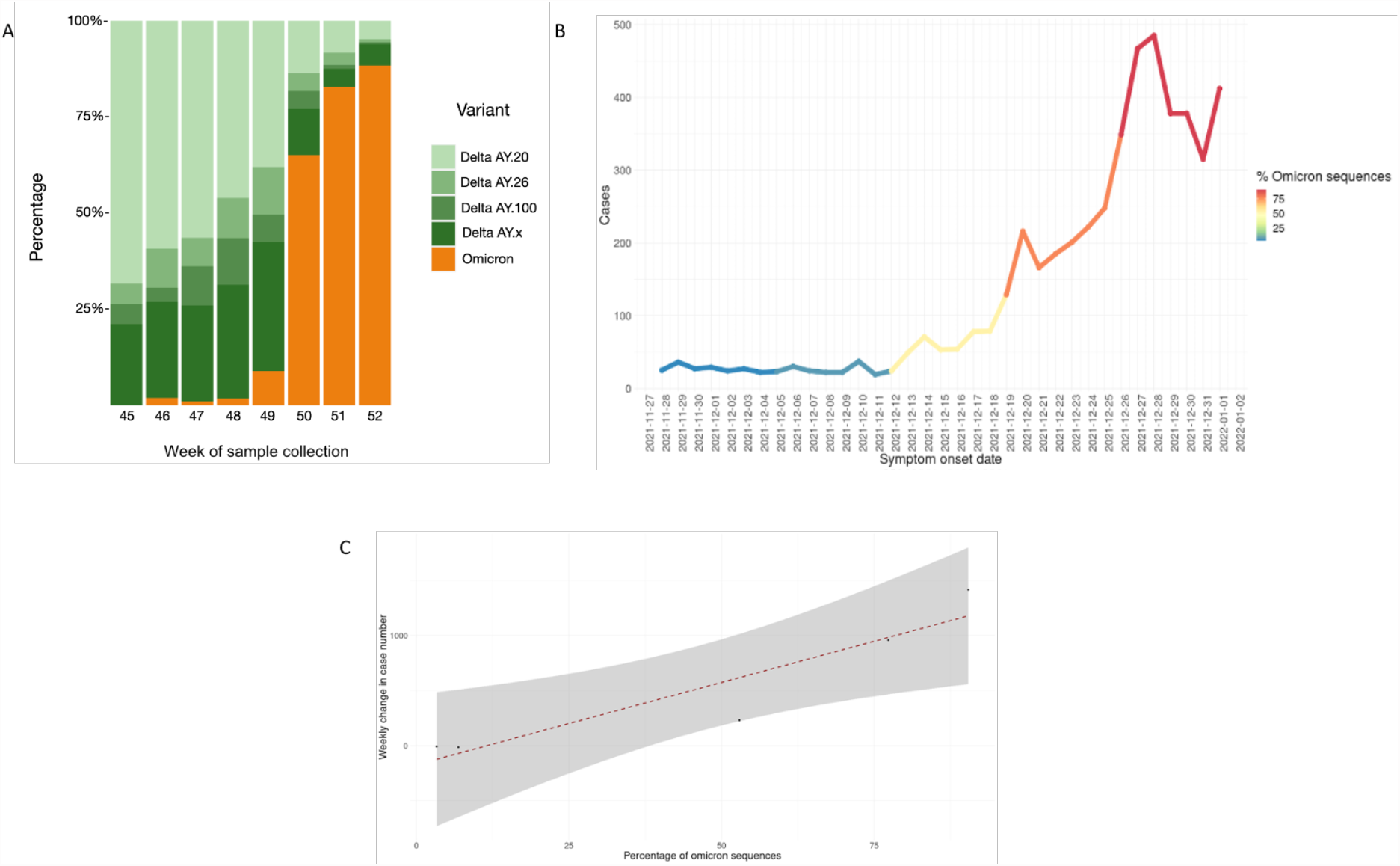
A. Prevalence of the Omicron variant in Mexico City from 16 November to 31 December 2021 (week 45 to week 52). B. % of Omicron variant cases and its symptom onset date. C. Correlation plot between (weekly) growth rate of the epidemic curve and the proportion of Omicron variant.

Never before had a variant of concern (VOC) displaced other variants so quickly in Mexico City, in 2021 variant B.1.1.519 was totally displaced by Alfa, Gamma and Delta in four months [5], while the Omicron variant displaced Delta in only one month. There was a correlation between the growth of the epidemic curve and the proportion of Omicron (Figure 2C). The increase in SARS-CoV-2-positive cases began on December 29, 2021, in Mexico City in four waves, a month and a half after the first detection of Omicron by sequencing, which speaks to the importance of SARS-CoV-2 genomic surveillance programs for public health decision making [20].

### Omicron genetic background

Of the 783 Omicron genomes sequenced from November 16, 2021, to December 31, 2021, in Mexico City, the region coding for the spike protein contains 50 nonsynonymous mutations, 19 of which are located in the receptor binding domain RBD). Figure 3 shows the most frequent amino acid substitutions in the RBD (G339D, R346K, K417N, S371 L, S373P, S375F, N440K, Q498R and N501Y). N501Y is present in Alpha, Beta and Gamma variants, and this substitution increases binding affinity to angiotensin-converting enzyme receptor 2 (ACE2), playing an essential role in the higher rate of transmission of SARS-CoV-2 variants [21]. K417N/T is present in alpha and gamma variants, and both mutations facilitate immune escape for monoclonal antibodies (bamlanivimab/LY-CoV555) [22], escape from neutralization by convalescent plasma [23], and escape by sera from BNT162b2-vaccinated individuals [24]. These results indicate that Omicron has a significant immune escape from an existing protection established by virus infection or vaccination, most likely due to the accumulation of the mutations described above [25,26].

**Figure 3.**
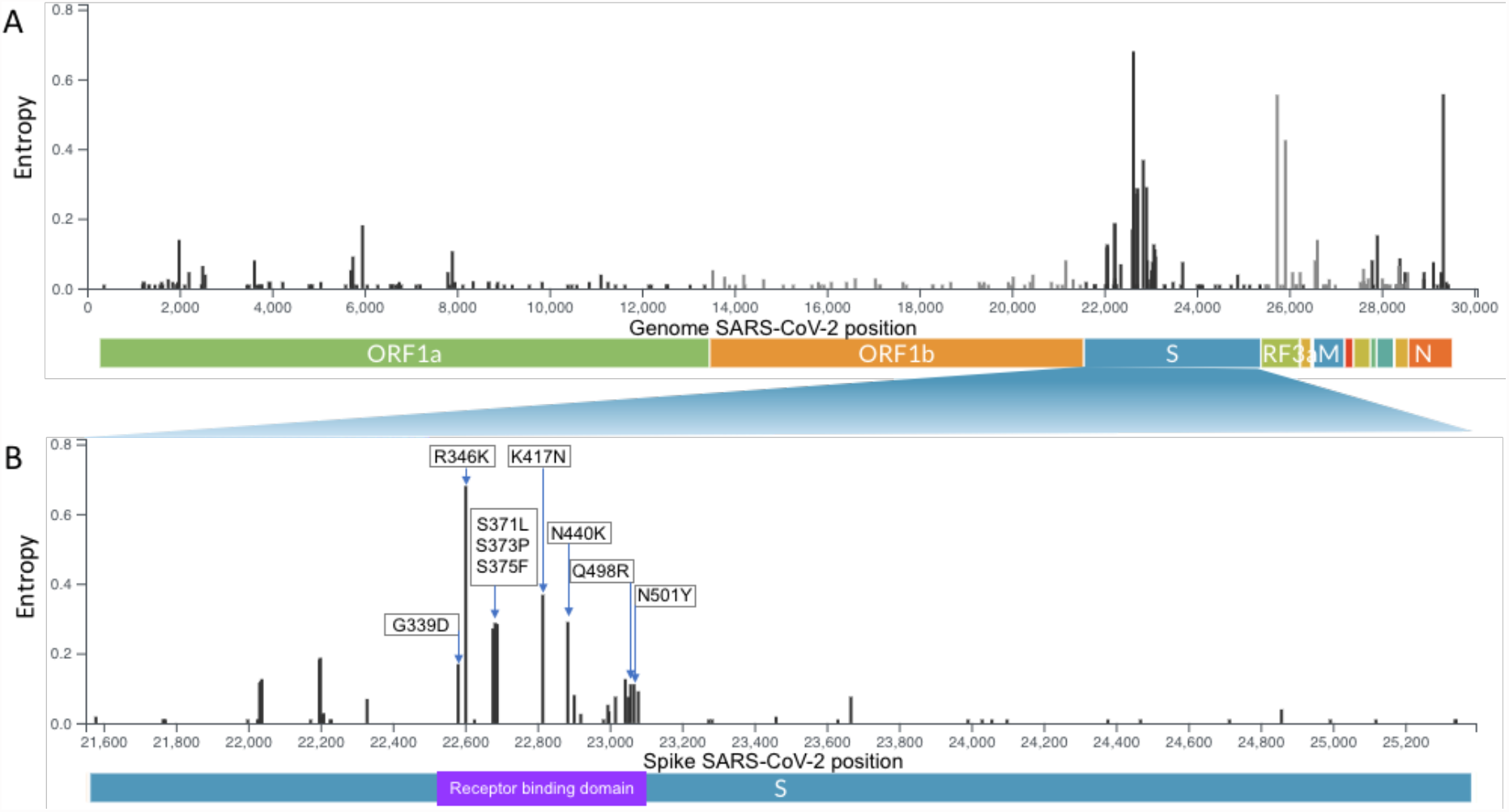
A. Genome map of the SARS-CoV-2 Omicron variant with the most representative amino acid substitutions in 783 Omicron genomes in Mexico City with genome coverage >95%. Whole genome SARS-CoV-2. B. Spike protein with receptor binding domain. The y-axis corresponds to the entropy calculated by the nextclade tool.

We performed a hierarchical clustering of the prevalence of the most frequent substitutions in the spike protein in the top 3 countries that generated the most Omicron genomes per continent. This analysis shows that the prevalence in substitutions is similar between Mexico City and Mexico no-Mexico City, Germany and Brazil since they clustered together.

We found that the substitutions R346K, K417N and N440K were more prevalent in Mexico City, Mexico no-Mexico City, USA and Japan (42-52%). In addition, a group of five substitutions (A67V, del69/70, T95I, G142D, del143/145) had the highest prevalence values in all countries analyzed (92-100%), except in the United Kingdom and India (60-71%).

On the other hand, the prevalence of eleven substitutions prevalence (S371L(S371L, S373P, S375F, S477N, T478K, E484A, Q493R, G496S, Q498R, N501Y and Y505H) was lower in Mexico City and Mexico no-Mexico City, Israel, United Kingdom and India (32-70%) than in Australia, Canada, Japan, and the USA (91-100%) (Figure 4).

**Figure 4.**
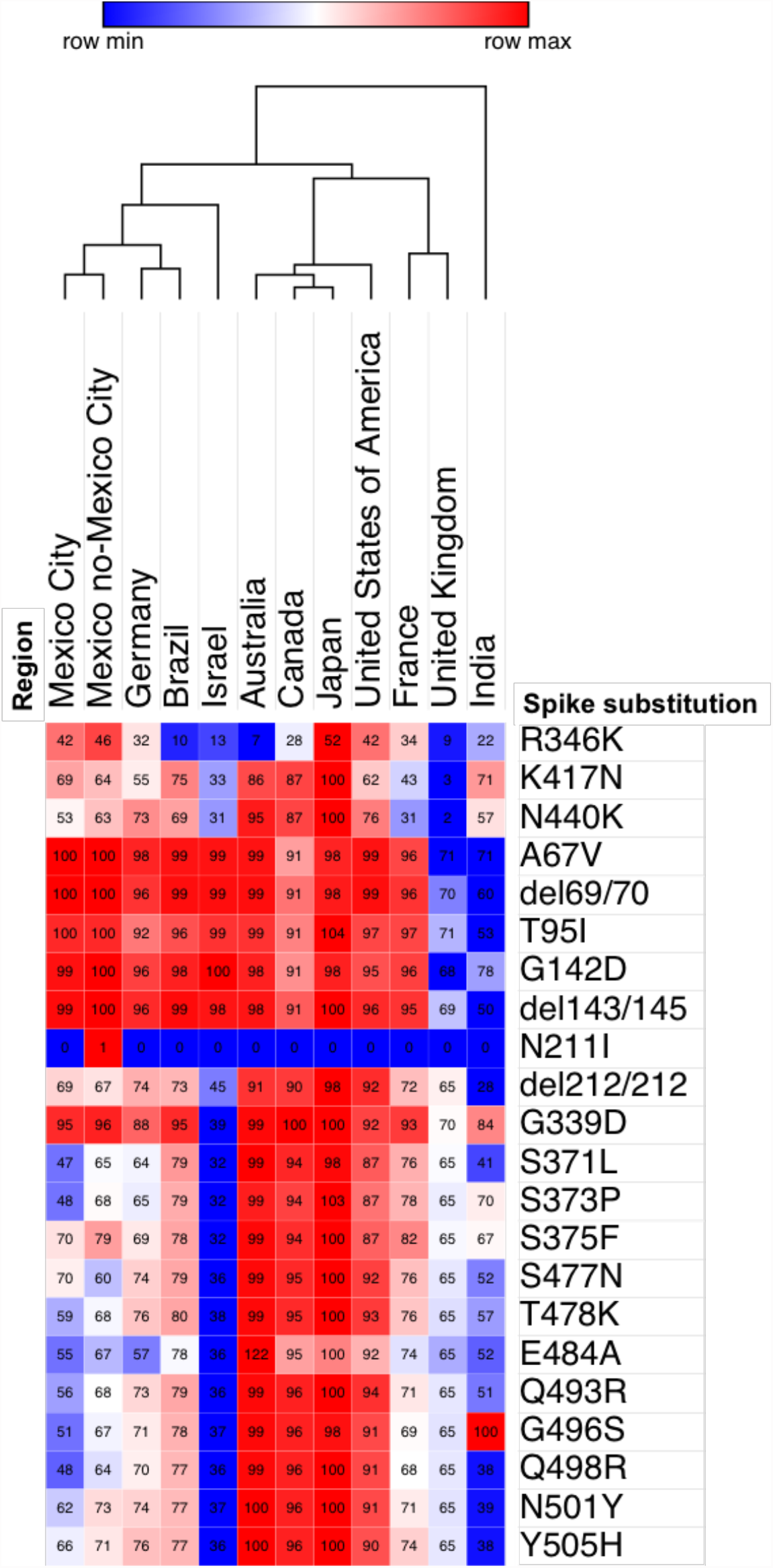
Hierarchical clustering of the prevalence of amino acid substitutions in spike among different countries in samples collected from 16 November to 31 December 2021 obtained from GISAID. We use one minus the Pearson correlation and clustering by region and the average as the linkage method. The scale represents the prevalence by each substitution in countries. Each cell contains the prevalence of the substitution as a percentage.

### Prevalence of R346K substitution

The prevalence of R346K was 42% in Mexico City and 46% in the rest of the country; other countries with high prevalence were the USA (42%) and Japan (52%). Interestingly, only the samples with R346K in México City clustered in a monophyletic branch in a phylogenetic analysis (Supplementary Figure 1). R346K was presented for the first time in the Mu variant B.1.621 [27] and is now present in the Omicron variant. This substitution has clinical importance in the therapy of COVID-19. Neutralization studies on the B.1.1.529+R346K pseudovirus showed that 18 of the 19 monoclonal antibodies (mAbs) tested lost neutralizing activity completely or partially [28]. There are mAB-based therapies that have demonstrated efficacy in neutralizing Omicron+R346K, such as STI-9167 from Sorrento Therapeutics [29], Sotrovimab by GSK and Vir Biotechnology [30].

### Haplotype network

Virus evolution can be represented by an haplotype network. Any node in an haplotype network represents a virus haplotype, i.e. an unique combination of alleles present in at least one patient sample (Supplementary tables 1 and 2 and Supplementary figure 2). The weight of an edge represents the number of mutations between the two connected haplotypes, i.e. the number of mutations required to transit from one haplotype to another. The final haplotype network is the maximum parsimony representation of virus evolution as the total weight of the network is minimized during the construction of an haplotype network.

We build an haplotype network from Omicron SARS-CoV-2 sequences from Mexico City (Figure 5). Some haplotypes derive in a big number of descendent haplotypes producing star-like structures in the network; this phenomenon has been seen in previously reported haplotype networks from the SARS-CoV-2 and other viruses [31,32]. The big nodes represent common haplotypes as they are observed in a big number of patients. The color is proportional to the fraction of patient samples that were fully vaccinated from each haplotype. The white nodes belong to haplotypes with 100% non vaccinated patients and dark purple nodes belong to haplotypes with 100% full vaccinated patients. The red-arrowed haplotypes correspond to common haplotypes, as they are present in 6 and 4 patients, respectively, all of them fully vaccinated, further investigation would be required to understand this relationship. There are 64 haplotypes composed of solely fully vaccinated patients, 32 of them (50%) correspond to internal nodes, by other hand there are 21 haplotypes composed of solely not vaccinated patients, similarly 11 of them (0.53%) correspond to internal nodes. Internal nodes belong to haplotypes that diversified into other virus haplotypes. We observed that most haplotypes are present in only one patient making any interpretation difficult. Moreover in previous studies only marginal differences in haplotype distributions have been observed between low and high vaccination rate countries [33].

**Figure 5.**
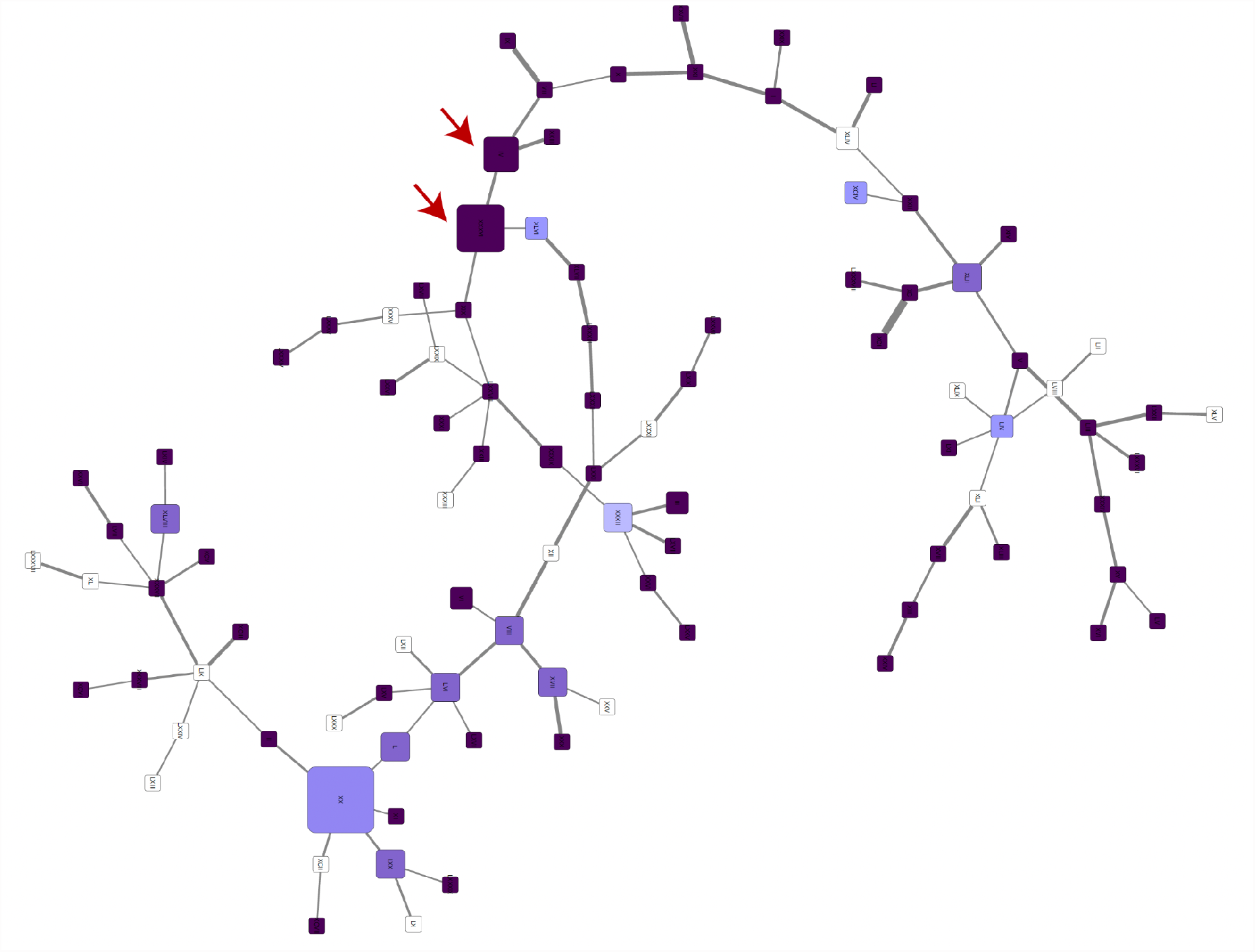
Haplotype network from all Omicron SARS-CoV-2 sequences from Mexico City. The size of each node is proportional to the number of samples that belong to that haplotype, and the color represents the fraction of samples that were fully vaccinated. The width of the lines is proportional to the number of mutations between two haplotypes.

### Epidemiology and clinical associations

As previously mentioned, we observed an increase in the prevalence of the Omicron variant over the previously predominant delta variant in the span of ∼1 month. During this transition, basic population descriptors (as seen in Figure 6) of the infected population, such as age distribution, sex distribution, number of reported comorbidities and number of reported symptoms at the time of diagnosis, remained virtually unchanged, with no significant differences between the variant groups (see Table 1).

**Table 1:** p values of the comparison (Fisher’s exact test) between the delta and Omicron variants.

| Variable | p.value |
| --- | --- |
| Sex | 0.6414 |
| Vax | 0.3285 |
| Hospitalization status | 0.1692 |
| Age | 0.1801 |
| Number of comorbidities | 0.7300 |
| Number of symptoms | 0.8343 |

\*

**Figure 6.**
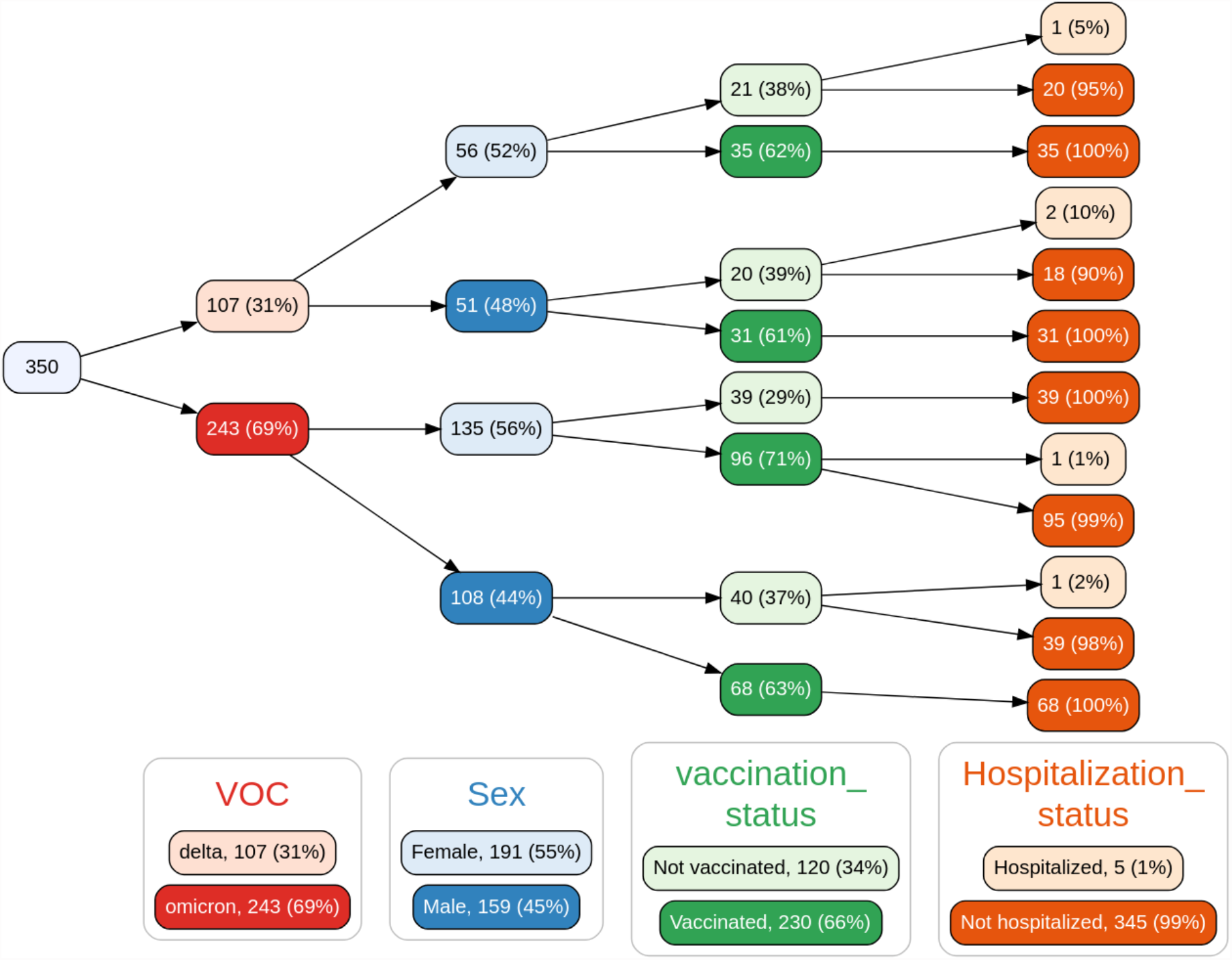
Variable tree with the spread of samples based on variant of concern (VOC), sex, vaccination status and hospitalization status. Not shown: a single sample in the delta->male->not vaccinated->hospitalized branch that is the only sequenced case deceased in the analysis period.

In terms of vaccination, our cohort was composed of 66% vaccinated individuals. It should be noted that in this cohort of sequenced samples, only five patients required hospitalization, and only one death was reported on the last day of the analysis period. Due to this reduced number of adverse outcomes, our analyses are not able to test whether there is an association between the Omicron variant and hospitalization or death outcomes in the analysis period, much less to model the effect of vaccination status as other groups have [34]. We should emphasize that the data analyzed in this work, which are essentially early-stage variant introduction data, neither support nor reject the notion of a difference in the likelihood of adverse outcomes associated with the Omicron variant becoming the dominant variant, particularly in the context of policymaking and mitigation strategies; the intrinsic severity of Omicron is still an open question that should be addressed with great care [35].

We assessed whether any of the measured comorbidities that are routinely reported in the Mexican COVID-19 case recording protocol were differently represented in the Omicron or delta populations. The logistic regression model showed that none of these comorbidities were associated with a specific variant. These results suggest that risk assessment based on comorbidities and associated clinical risk predictors currently used [4,36] may remain valid for the Omicron variant.

In terms of symptomatology, a logistic regression model (see table 2) showed two symptoms as significant to differentiate between the delta and Omicron variants. These are dysgeusia (distortion of taste) and odynophagia (pain with swallowing). The first one is more associated with the delta variant, while the second one is more associated with the Omicron variant. In Figure 7, we illustrate this association by showing the percentage of all recorded cases in Mexico City (regardless of sequencing status); a drop in dysgeusia is clearly observable as Omicron becomes more dominant. While there have been reports on the media and in the recent medical literature [37] of changes in the symptomatology associated with the Omicron variant, the modest results of our regression model suggest that a diagnosis of COVID-19 caused by the Omicron variant through symptoms alone may be unfeasible, highlighting the need for proper genomic surveillance. However, monitoring signals such as the drop in reports in dysgeusia may provide early warning signs of the introduction of the variant to new populations and trigger assessment using proper genomic tools.

**Table 2:** Logistic regression -variants as a function of symptoms (delta variant as baseline). Predictors with p < 0.05 are shown.

| Term | odds ratio | p.value |
| --- | --- | --- |
| Intercept | 2.01 | 0.0021 |
| Odynophagia | 2.10 | 0.0090 |
| Dysgeusia | 0.103 | 0.0009 |

**Figure 7:**
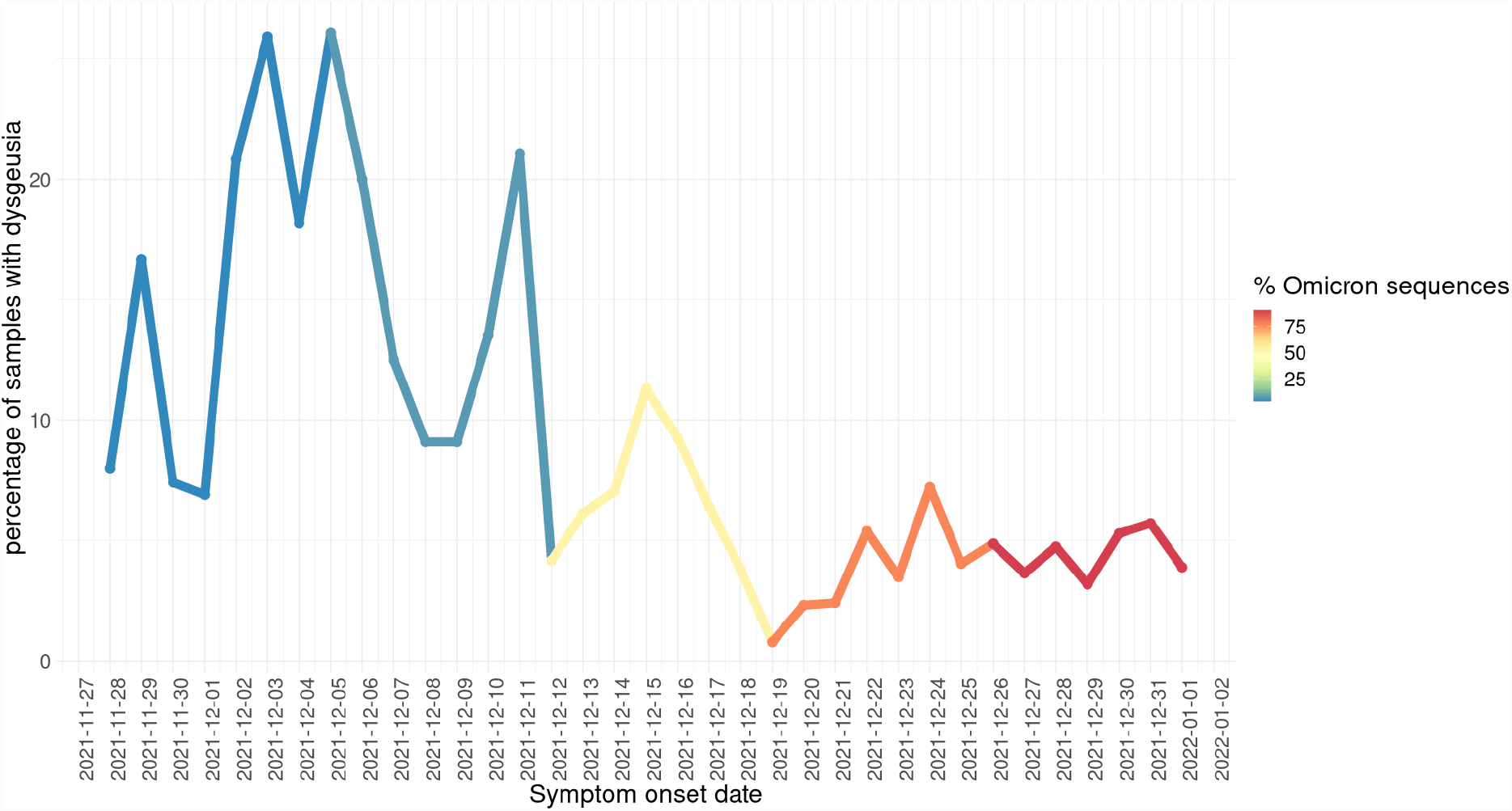
Daily fraction of all COVID-19-positive cases detected in Mexico City that reported dysgeusia as a symptom (by symptom onset date). Line color indicates the (weekly) percentage of sequenced samples identified as Omicron detected by genomic surveillance.

We also analyzed the difference between Ct distribution for the samples classified as either delta or Omicron for each of the markers amplified during the qRT-PCR test. The distributions are shown in Figure 8. We observed a sharp difference on the amplification of the spike gene complying with the detection kit manufacturer’s observation of a dropout of the S-gene target associated with the Omicron SARS-CoV-2 variant. This dropout was first identified in the Alpha variant. It is caused by the 69-70del mutation of the S gene that interferes with the amplification of the S-gene target. Our results suggest that a qRT-PCR test could be an initial and efficient approach to propose the variant classification of a patient sample at least at this moment of the pandemia where practically all samples belong to either delta or Omicron variants. Besides, epidemiological surveillance and monitoring are important to rapidly detect unexpected behaviors during qRT-PCR amplification that could be associated with the emergence of new variants.

**Figure 8.**
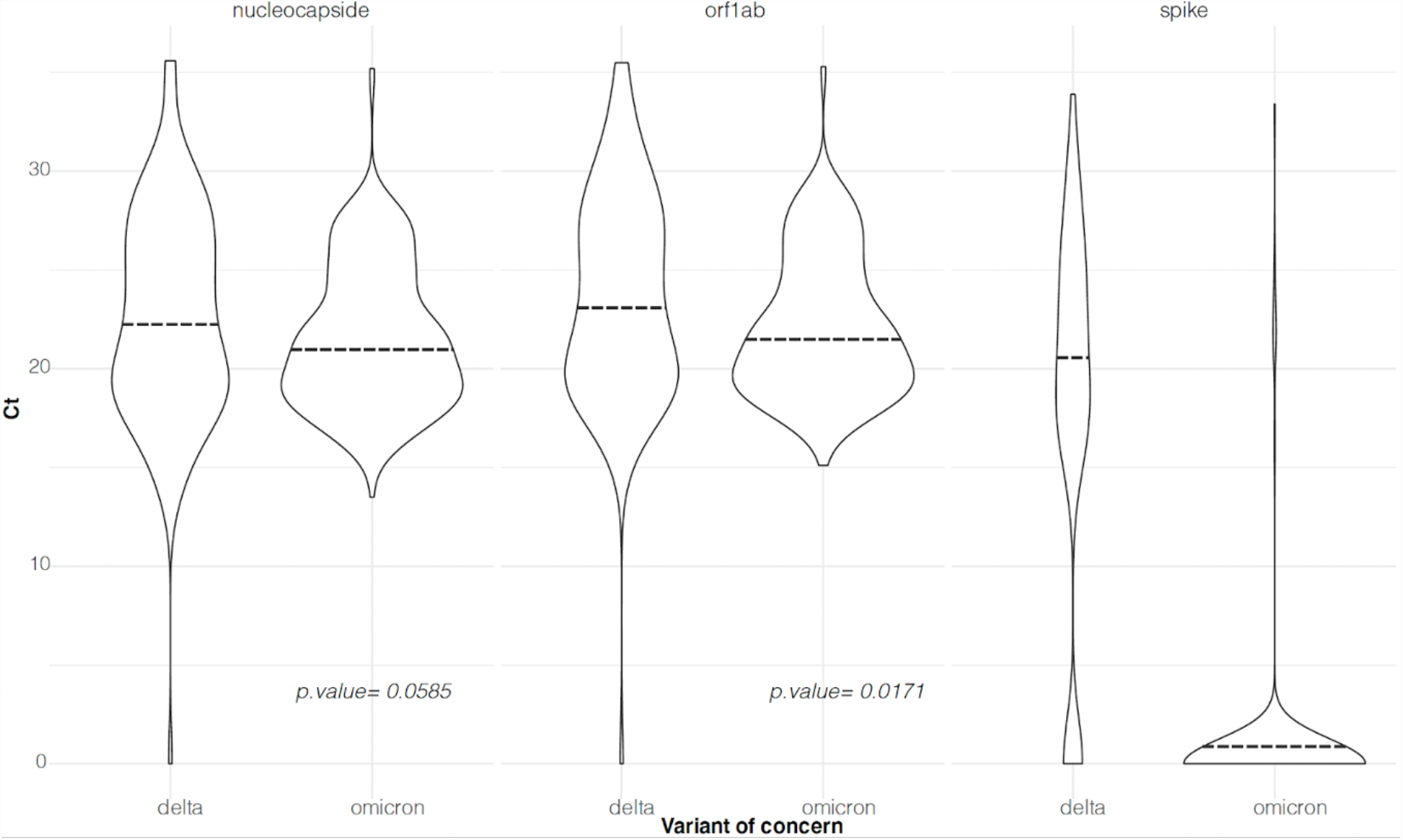
Ct distributions for each marker used during the qRT-PCR test for delta and Omicron SARS-CoV-2 samples. A two-sample t-test was performed for each marker. The corresponding p value is shown in each case. Panel A, distribution of Cts observed during nucleocapside target amplification. Panel B, distribution of Cts observed during orf1ab target amplification. Panel C, distribution of Cts observed during spike target amplification. The dashed lines represent the mean for each distribution.

We performed a t-test on the nucleocapsid and orf1ab Ct distributions to compare the Ct mean between the delta and Omicron variant. We found a non-significant difference (p value = 0.0585) in the case of the nucleocapsid marker and a significant difference in the case of the orf1ab marker (p value = 0.0171). Although viral load and Ct cannot be directly correlated [38,39], the possible clinical significance of the observed change remains to be investigated.

## Conclusion

Mexico City is the most populated city in Mexico as well as the political and economic center of the country. It is also one of the largest tourist and commercial entry points. Epidemiological surveillance in high-movement urban regions such as Mexico City is important to timely detect the appearance and propagation of new SARS-CoV-2 variants even weeks before the highest number of cases is reported. In this study, we reported no significant clinical differences between the populations infected by the delta or Omicron variants; however, we reported symptomatology differences associated with each of these variants. Therefore, it is important to continue monitoring the pandemic behavior to detect patterns that could inform public health decisions and guide decision-makers. As the pandemic is a highly dynamic phenomenon, it is important to continue genomic surveillance to detect the propagation of any mutation that could affect treatment selection or effectiveness, as well as changes in the clinical features and public health indicators associated with the spread of the virus.

## Data Availability

All data is available on GISAID website https://www.gisaid.org/

