## Supplementary Figures for "Early genomic, epidemiological, and clinical description of the SARS-CoV-2 Omicron variant in Mexico City"

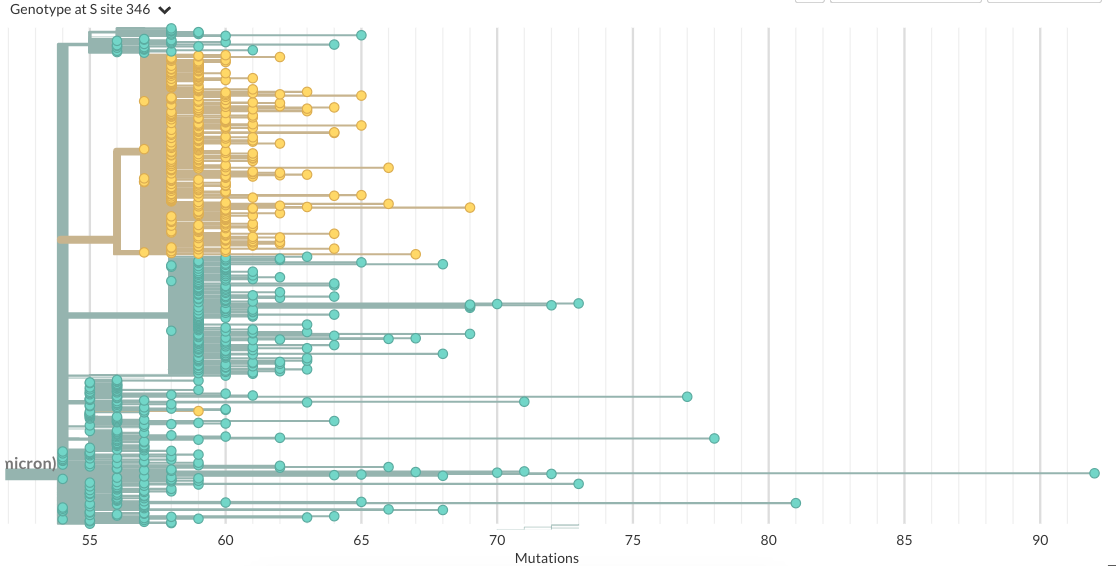

**Supplementary Figure 2.** *Phylogeny of omicron samples in Mexico City, samples with R346K substitution clustering in a monophyletic group.*

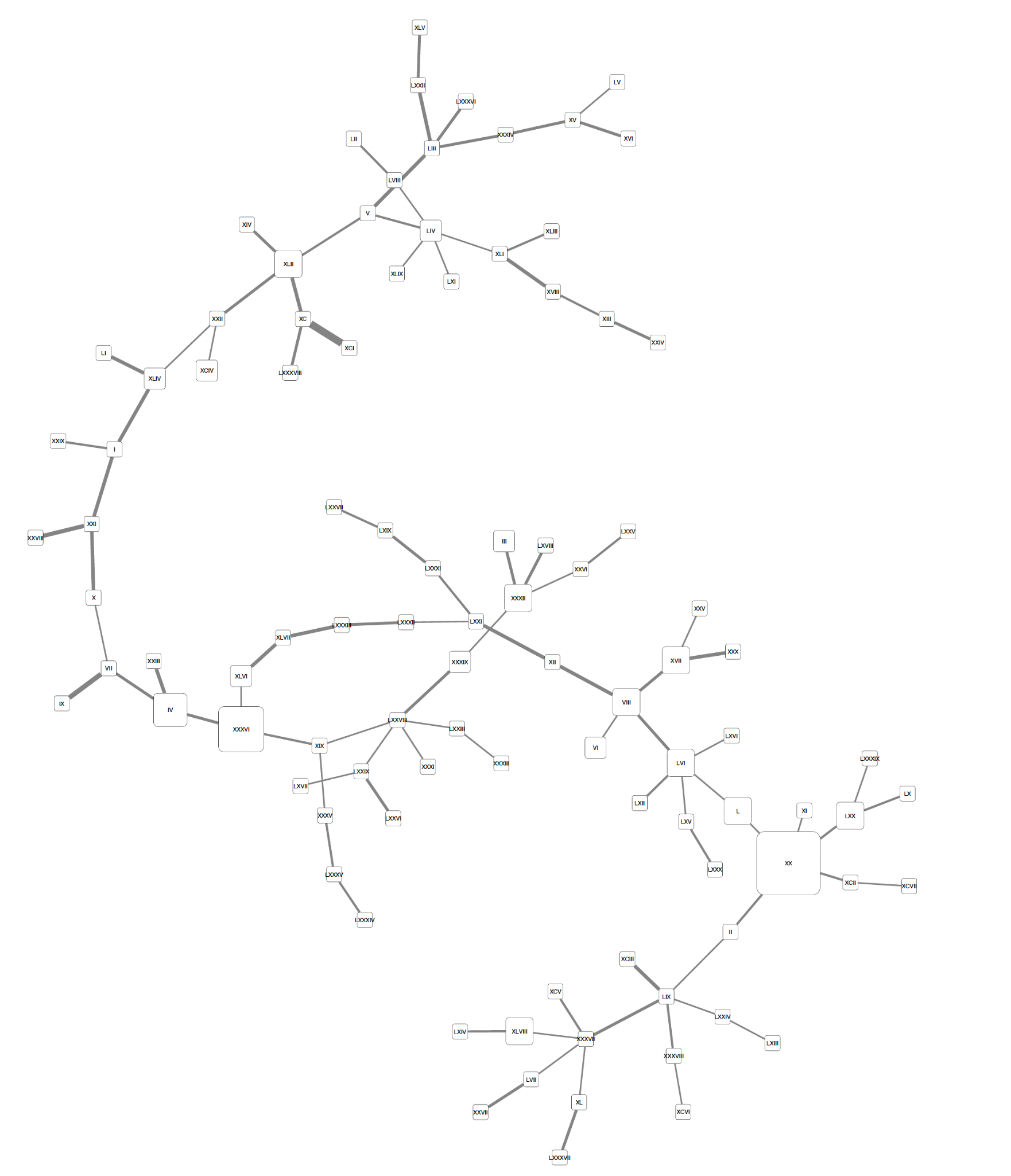

**Supplementary Figure 2.** *Haplotype network from all omicron SARS-CoV-2 sequences from Mexico City. The size of each node is proportional to the number of samples that belong to that haplotype. The width of the lines is proportional to the number of mutations between two haplotypes. This figure is the same as FigureX on the main text but rotated so the haplotype numbers are visible.*

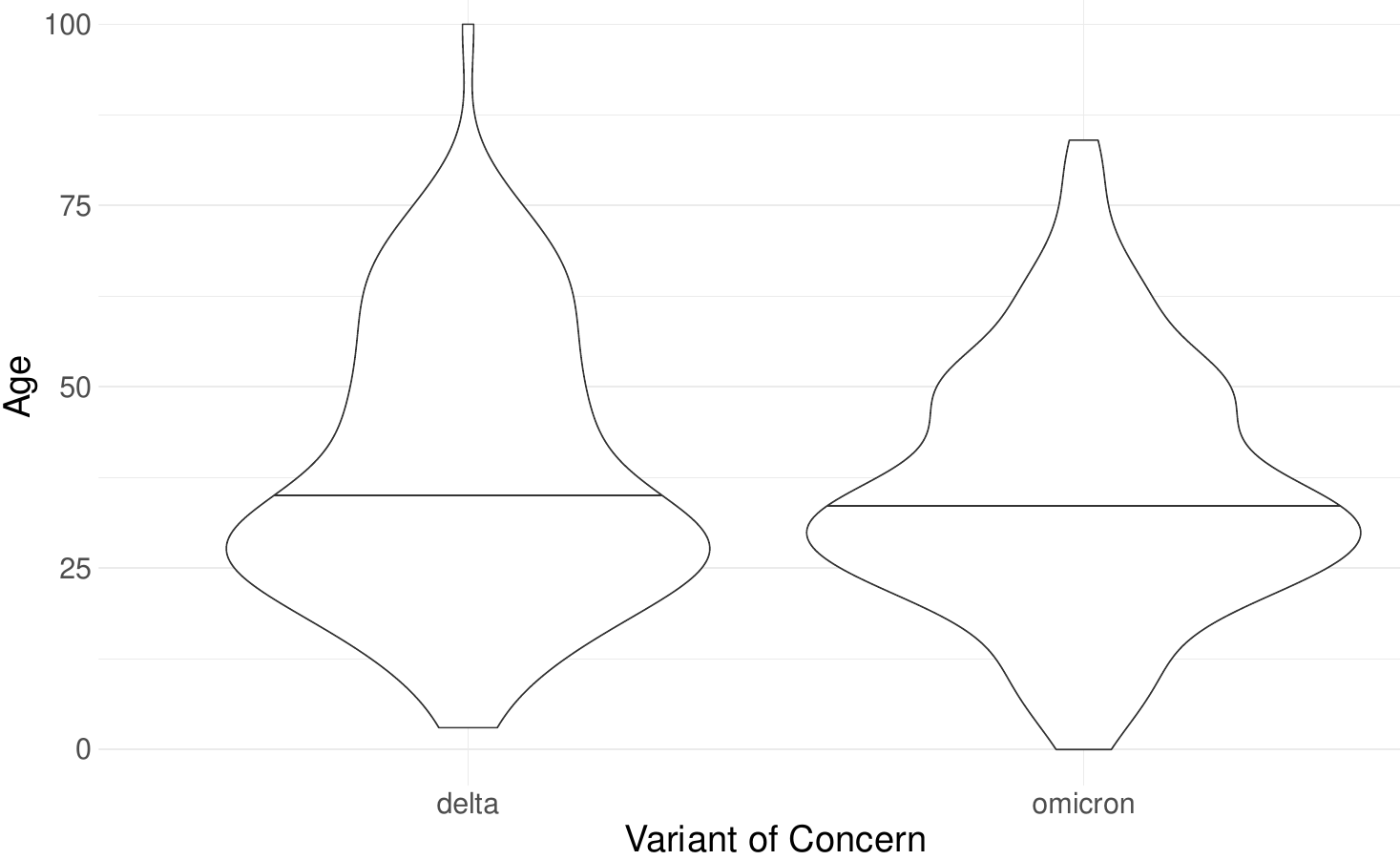

***Supplementary Figure 3:*** *Violin plot of age distribution by VOC, showing no significant differences between groups.*

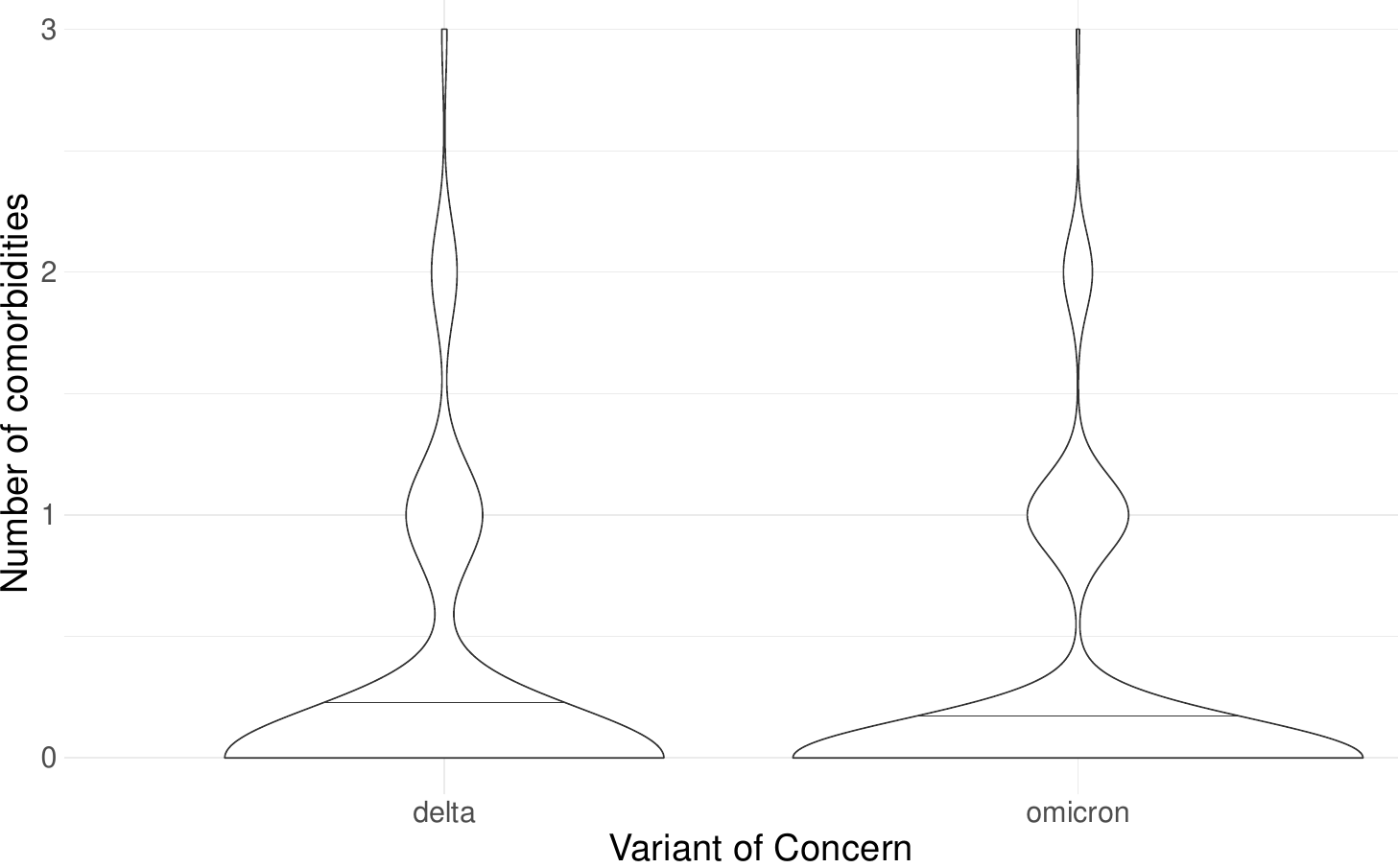

***Supplementary Figure 4:*** *Violin plot of number of comorbidities distribution by VOC, showing no significant differences between groups.*

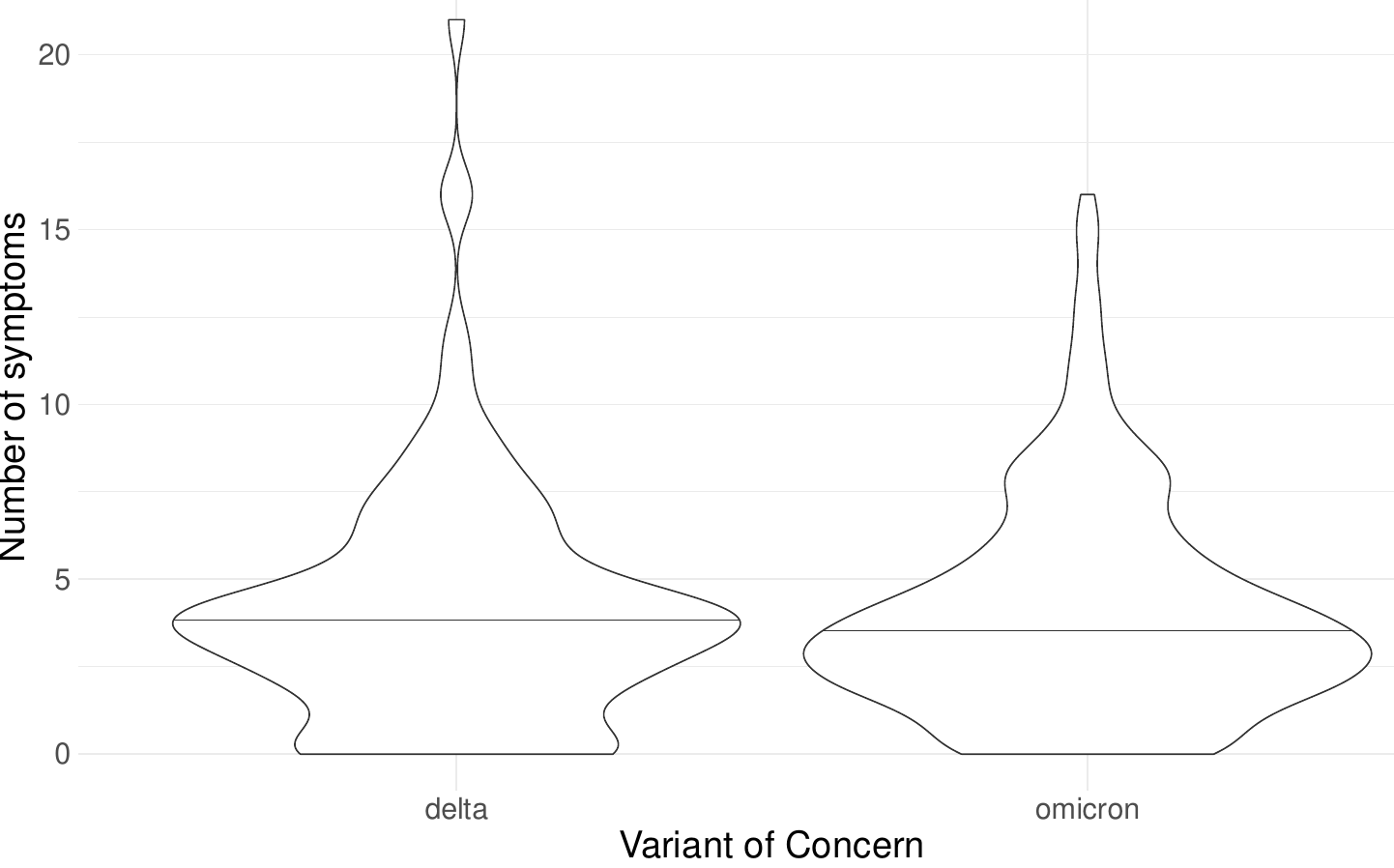

**b**

***Supplementary Figure 5:*** *Violin plot of symptoms number distribution by VOC,, showing no significant differences between groups.*
